# Physical and brain frailty in stroke: shared occurrence and outcomes. A cohort study

**DOI:** 10.1101/2023.02.15.23286006

**Authors:** M Taylor-Rowan, M Hafdi, B Drozdowska, E Elliott, J Wardlaw, T Quinn

**Affiliations:** Institute of Health and Wellbeing, University of Glasgow, UK; Department of Neurology, Amsterdam UMC, University of Amsterdam, Netherlands; Institute of Cardiovascular and Metabolic Sciences, University of Glasgow, UK; Centre for Clinical Brain Sciences, Edinburgh Center in the UK Dementia Research Institute, University of Edinburgh, UK

**Keywords:** Frailty, cognition, dementia, stroke

## Abstract

**Background:** There is increasing interest in the concept of frailty in stroke, including both physical frailty and imaging-evidence of brain frailty. We aimed to 1)establish concurrent validity of a brain frailty measurement against traditional measures of physical and global frailty 2)establish prevalence of brain frailty in stroke survivors with and without pre-existing frailty 3)establish the predictive validity of measures of physical, global, and brain frailty against long-term cognitive outcomes.

**Methods:** We included consecutively admitted stroke or transient ischaemic attack (TIA) survivors from participating stroke centres. Baseline CT scans were used to identify leukoaraiosis, atrophy, or old infarcts; these were then combined to generate an overall brain frailty score for each participant on a scale ranging from 0-3 (3=higher brain frailty). Global frailty was measured using Rockwood frailty index, and physical frailty using a Fried frailty screening tool. Presence of major or minor neurocognitive disorder at 18-months following stroke was established via a multicomponent assessment. We assessed the concurrent validity of brain frailty and frailty scales via Spearman’s rank correlation. Prevalence of brain frailty was established based upon observed percentages within groups defined by frailty status (robust, pre-frail, frail). We conducted multivariable logistic regression analyses to evaluate association between brain, global, and physical frailty with 18-month cognitive impairment.

**Results:** Three-hundred-forty-one stroke survivors participated. Brain frailty was weakly correlated with Rockwood frailty (Rho:0.336;p<0.001) and with Fried frailty (Rho:0.230;p<0.001). Three-quarters of people who were frail had moderate-severe brain frailty and prevalence increased according to frailty status. Brain frailty (OR:1.64,95%CI=1.17-2.32), Rockwood frailty (OR:1.05,95%CI=1.02-1.08) and Fried frailty (OR:1.93,95%CI=1.39-2.67) were each independently predictive of cognitive impairment at 18 months following stroke. Fried frailty was predictive independent of age, sex, stroke severity, education, baseline cognitive test performance, and brain frailty status (OR:1.49,95%CI=1.03-2.18)

**Conclusions:** Physical and brain frailty are separate concepts, although they frequently co-exist. Both are associated with adverse cognitive outcomes and physical frailty remains important when assessing cognitive outcomes.

## Introduction

Frailty is a highly prevalent condition amongst stroke survivors, existing in as many as 28% of patients upon admission to the acute stroke unit.^1, 2^ Frailty assessment is increasingly being incorporated into acute care pathways but has yet to be routinely adopted in stroke.^2^ Evidence suggests there is prognostic value in assessing for frailty in stroke: delirium, mortality and post-stroke cognitive outcomes are all associated with frailty status.^3, 4^

Frailty can be characterised as a state of multisystem physiological vulnerability with diminished capacity to manage external stressors.^5^ It is typically assessed by measuring symptoms of physical frailty, as developed by Fried^6^, or via a more global measure of frailty that encapsulates a combination of physical and neuropsychological conditions, as developed by Rockwood.^7^

Following stroke, evidence of pre-existing brain frailty, i.e. damage visible on a brain scan at the time of the stroke but that must have been present prior to the stroke, may also provide valuable prognostic information.^8^ In general, brain frailty is a state of reduced neurophysiological reserve and may predispose people to poor cognitive outcomes.^9^ Functional evidence of brain frailty may be underpinned by evidence of diffuse or focal vascular or neurodegenerative brain damage on CT or MR brain imaging. Recently, Appleton^8^ tested a scale for measuring brain imaging-based brain frailty^10^ that involves routinely observable neuroimaging markers (leukoaraiosis, cerebral atrophy, and old infarcts). However, the scale has not yet been validated directly against traditional functional measures of frailty and we do not know the extent to which imaging-derived brain frailty co-occurs with physical or global measures of functional frailty.

Traditional measures of functional frailty such as Fried or Rockwood tend to be considered in isolation from directly observed imaging-derived brain frailty despite increasing evidence indicating they may be associated.^11, 12^ If traditional measures of frailty are synonymous with brain frailty, then measuring each construct separately is inefficient. On the other hand, if they are distinct, establishing their independent predictive value could help improve clinical decision making.

## Aims

Our study had 3 main aims:

1. to validate a measure of imaging-derived brain frailty against traditional (physical and global functional) measures of frailty.
2. to establish the prevalence of brain frailty in those who are/are not frail (as defined according to each traditional measure).
3. to examine the association between pre-stroke frailty and pre-stroke brain frailty with long-term cognitive impairment and dementia following stroke.

## Methods

We followed STROBE (Strengthening the Reporting of Observational Studies in Epidemiology) guidelines^13^ for reporting. This is a sub study of the APPLE (Assessing Psychological Problems in stroke: A longitudinal Evaluation; research registry ID:1018) project—a multicentre, prospective longitudinal cohort study embedded within the UK National Health Service. A summary of our methodology is provided below. Comprehensive details are available in the study protocol.^14^ Ethical approval was approved by the Scotland A Research Ethics committee and obtained for all participating sites (REC number:16/SS/0105).

### Setting

APPLE recruited consecutively admitted stroke and transient ischaemic attack (TIA) survivors admitted to participating acute stroke centres (November 2016 to February 2019) with no exclusions based on age, stroke-type, stroke-severity, or comorbidity. This sub-study was restricted to sites with CT brain raw images available for analysis.

### Brain frailty assessment

Baseline CT brain scans were performed for all participants as part of routine acute care. We adopted an approach developed in IST-3^10^ and further refined for application,^8^ to generate a brain frailty score for each participant using baseline CT scan images. A detailed description of this approach can be seen in the protocol.^15^ In brief, two trained assessors (MH; MT), who were blinded to clinical details, rated baseline CT scans using a set proforma^15^ and scored presence of leukoaraiosis, cerebral atrophy, and old infarcts. Scans were rated against a standard template **(supplementary materials Figure S1)**. ‘Modest’ leukoaraiosis, ‘modest’ atrophy (centrally and/or cortically), and presence of old focal cortical or subcortical vascular lesions were combined to generate a brain frailty score ranging from 0 (no brain frailty) to 3 (severe brain frailty).

### Frailty assessment

We measured frailty according to two of the most commonly employed frailty concepts: Rockwood’s ‘accumulated deficits’ global measure of frailty, and Fried’s ‘frailty phentoype’, which focuses on physical frailty.

Rockwood frailty was measured via a 32-item frailty index, (**supplementary materials Figure S2)** constructed according to recommended guidelines.^16^ A combination of participant medical records and self/informant reported functional measures^17^ and Independent Activities of Daily Living^18^ were used to identify physical, functional, cognitive, or psychological issues present before the stroke (informant reports were only used in cases where the participant was not able to complete a self-reported measure). Possible scores ranged from 0-100, with scores closer to 100 suggesting greater frailty. Participants were categorised as ‘robust’, ‘pre-frail’ and ‘frail’ using recommended cut-points of <8; 8-24; and >24, respectively.

Fried frailty was established via a self-report frailty screening tool.^19^ Where possible, informant reports were used when self-reported information was unavailable. The measure involves self-reported symptoms of physical frailty including exhaustion, unexplained weight loss, and low physical activity. Cut-points recommended by the scale developers were used to identify ‘frail’ and ‘non-frail’ participants.

### Outcome assessment

We employed a centrally adjudicated approach to establish presence of major and minor cognitive impairment at 18 months following stroke. Participants received a multi-domain cognitive assessment battery (details are proved in the APPLE protocol^14^) at baseline, 1 month, 6 months, 12 months, and 18 months. Performance on cognitive assessment was supplemented by evaluation of clinical case notes and other medical records up to an 18-month follow-up timeframe. Two assessors (medical and psychology backgrounds) independently evaluated the available data for each participant, and a consensus diagnosis, based on Diagnostic and Statistical Manual (DSM5) criteria,^20^ was established. Cognitive impairment diagnoses were applied conservatively; only formal diagnoses of major or minor neurocognitive impairment established via a medical professional, or consistent performance on cognitive testing that fell below standardised thresholds for impairment were adjudicated as impaired at 18-months. All adjudications were made without knowledge of participants’ frailty or brain frailty status.

### Statistical analyses

We conducted Spearman’s rank correlation to evaluate the concurrent validity of brain frailty relative to traditional frailty measures. The correlation coefficients were interpreted using conventional cut-points.^21^

Prevalence of brain frailty was established based upon observed percentages within the study population. Pre-specified cut-offs were applied to the Rockwood frailty index to differentiate ‘frail’, pre-frail and ‘non-frail’ participants. Fried frailty was established via dichotomisation at the recommended cut point.^19^ Confidence intervals were generated for each prevalence estimate.^22^

Multivariable logistic regression analysis was preformed to investigate the association between brain, physical and global measures of frailty with long-term cognitive impairment 18-months following stroke. Primary analysis (Model 1) controlled for age, sex, stroke severity and years in education. Brain frailty and Fried frailty were analysed on an ordinal scale, while Rockwood frailty was analysed on a linear scale.

We performed a sensitivity analysis restricted to participants with 18-month cognitive data only to ensure that our analysis was not overly influenced by adjudicated outcomes that relied more heavily on clinical case notes for 18-month diagnoses.

We conducted a series of subgroup analyses. In subgroup analysis 1, we excluded participants who had a known diagnosis of dementia at baseline in order to improve the clinical applicability of our analyses. In subgroup analysis 2, we additionally excluded people who had a TIA only.

We also conducted post hoc analyses to 1)explore if brain frailty had predictive power independent of physical or global frailty (Model 2), and vice-versa; 2)explore if brain, physical or global frailty had predictive power independent of baseline cognitive test scores—measured via the abbreviated mental test-plus (AMT-plus) (Model 3); 3)explore if brain frailty had predictive power over both baseline cognitive scores and frailty scores, or if physical or global measures of frailty had predictive power over both baseline cognitive scores and brain frailty scores (Model 4).

A univariate cox regression survival analysis was performed to investigate if there was a difference between brain, physical or global frailty status and likelihood of death during follow-up.

Regression analyses used a complete case approach. We conducted univariate logistic regression analyses to investigate differences between brain, physical or global frailty status and missing data. Statistical assumptions were checked for each analysis. SPSS version 27 (IBM) was used for all analyses.

## Results

Of 357 participants in APPLE, 341 were recruited from sites with CT brain data available.(**Figure1**,**Table1)** [Table1]

**Table 1.** Baseline study population characteristics

| Variable | Overall (N=341)* |
| --- | --- |
| *Age (Median; 25<sup>th</sup>-75<sup>th</sup> percentile)* | 71 (60-80) |
| *Sex Male (%)* | 188/341 (55.1%) |
| *Stroke-type (%)* |  |
| *Total Anterior Circulation Stroke* | 27/336 (8.0%) |
| *Partial Anterior Circulation Stroke* | 115/336 (34.2%) |
| *Lacunar Stroke* | 82/336 (24.4%) |
| *Posterior Circulation Stroke* | 66/336 (19.6%) |
| *Transient Ischaemic Attack* | 46/336 (13.7%) |
| *NIHSS (Mean; S.D.)* | 3.3 (4.4) |
| *Pre-stroke modified Rankin Scale (nn; %)* |  |
| 0-2 | 283/339 (83.4%) |
| 3-5 | 56/339 (16.5%) |
| *Vascular Disease (nn; %)* | 102/341 (29.9%) |
| *Heart Failure (nn; %)* | 26/341 (7.6%) |
| *Post-stroke Delirium (nn; %)* | 3/339 (0.9%) |
| *Post-stroke Aphasia (nn; %)* | 46/339 (13.6%) |
| *Previous stroke/TIA (nn; %)* | 86/341 (25.2%) |
| *Diabetes mellitus (nn; %)* | 82/341 (24.0%) |
| *Atrial Fibrillation (nn; %)* | 56/341 (16.4%) |
| *Alcohol Dependency (nn; %)* | 36/341 (10.6%) |
| *Education Years (Mean; SD)* | 11.9 (3.4) |

\*Some denominators ≠ 341 due to missing data

Three-hundred-thirty-two participants had baseline CT scan data available, 329 had Fried phenotype frailty scores available, and 339 had a Rockwood frailty index score. Thirty-three participants had missing clinical or demographic baseline data and 22 participants had missing baseline cognitive data. Follow-up cognitive test data was available for 266 (78%) participants at at least 1 time-point after baseline; 160 out of 308 (52%) surviving participants had 18-month cognitive test data available; secondary and primary care follow-up data were available for all participants (Figure 1).

**Figure 1:**
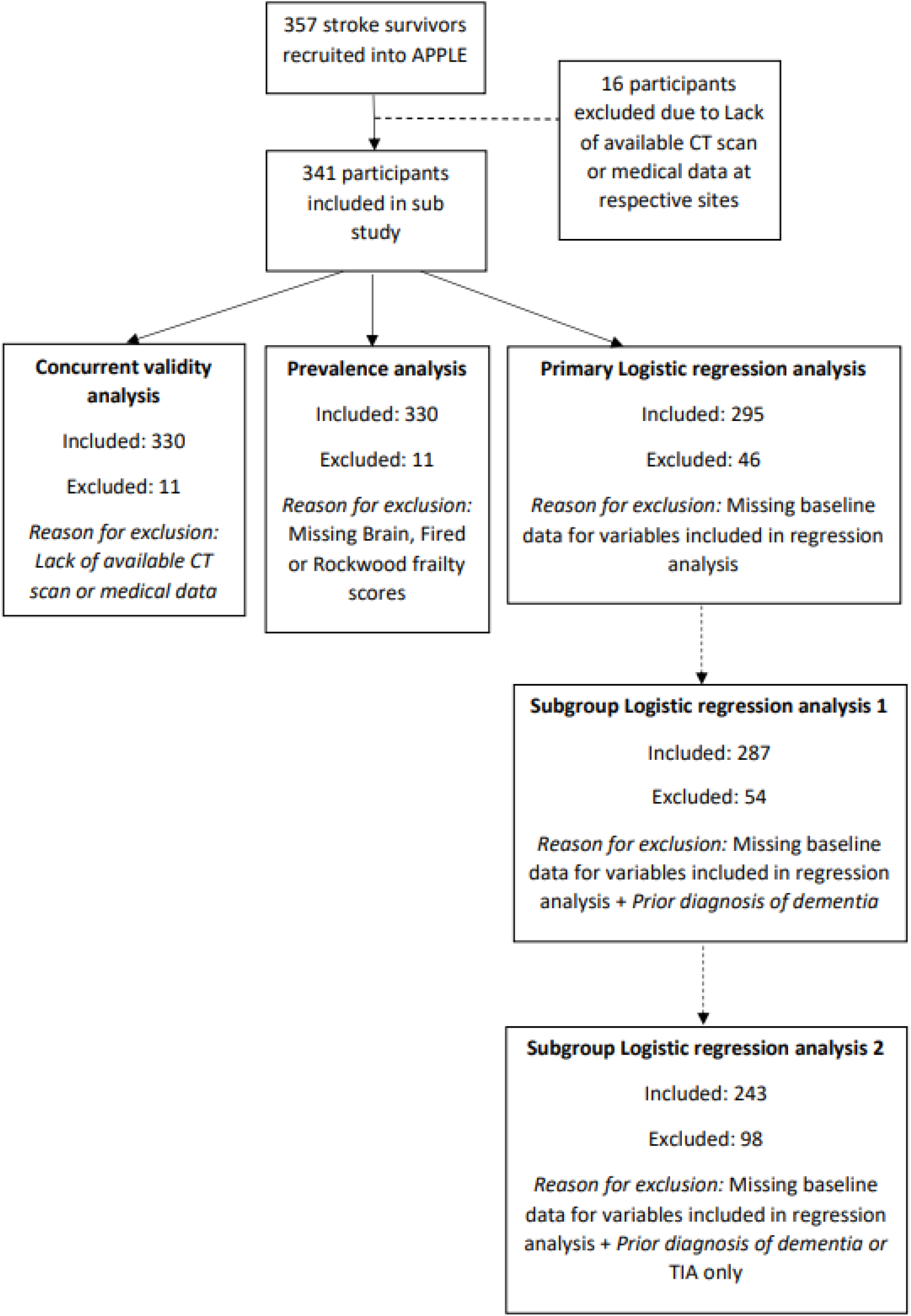
Flow chart of study attrition

Univariate regression analysis showed that brain frailty (OR:1.38, 95%CI=0.95-2.02) and Rockwood frailty scores (OR:1.02, 95%CI=0.98-1.06) were not associated with missing baseline data; but higher Fried frailty scores were associated with missing baseline data (OR:1.64,95%CI=1.12-2.41). Both brain frailty (OR:1.31, 95%CI=1.05-1.64), and Rockwood frailty (OR:1.04,95%CI=1.02-1.07) were associated with missing 18-month cognitive data but not Fried frailty scores (OR:1.15,95%CI=0.90-1.46).

Thirty-three (10%) participants died before end of study (18-months post-baseline); survival analysis indicated each point increase of brain frailty (HR:1.84, 95%CI=1.29-2.62), Rockwood frailty (HR:1.06, 95%CI=1.04-1.09), and Fried frailty (HR:1.59, 95%CI=1.14-2.22) significantly increased the risk of death before end of study.

### Concurrent validity of Brain frailty scale

Spearman’s rank suggested a significant, weak correlation between brain frailty and Rockwood frailty (rho:0.336; p<0.001). There was also a weak correlation between brain frailty and physical frailty (rho:0.230; p<0.001).(**Figure 2**)

**Figure 2:**
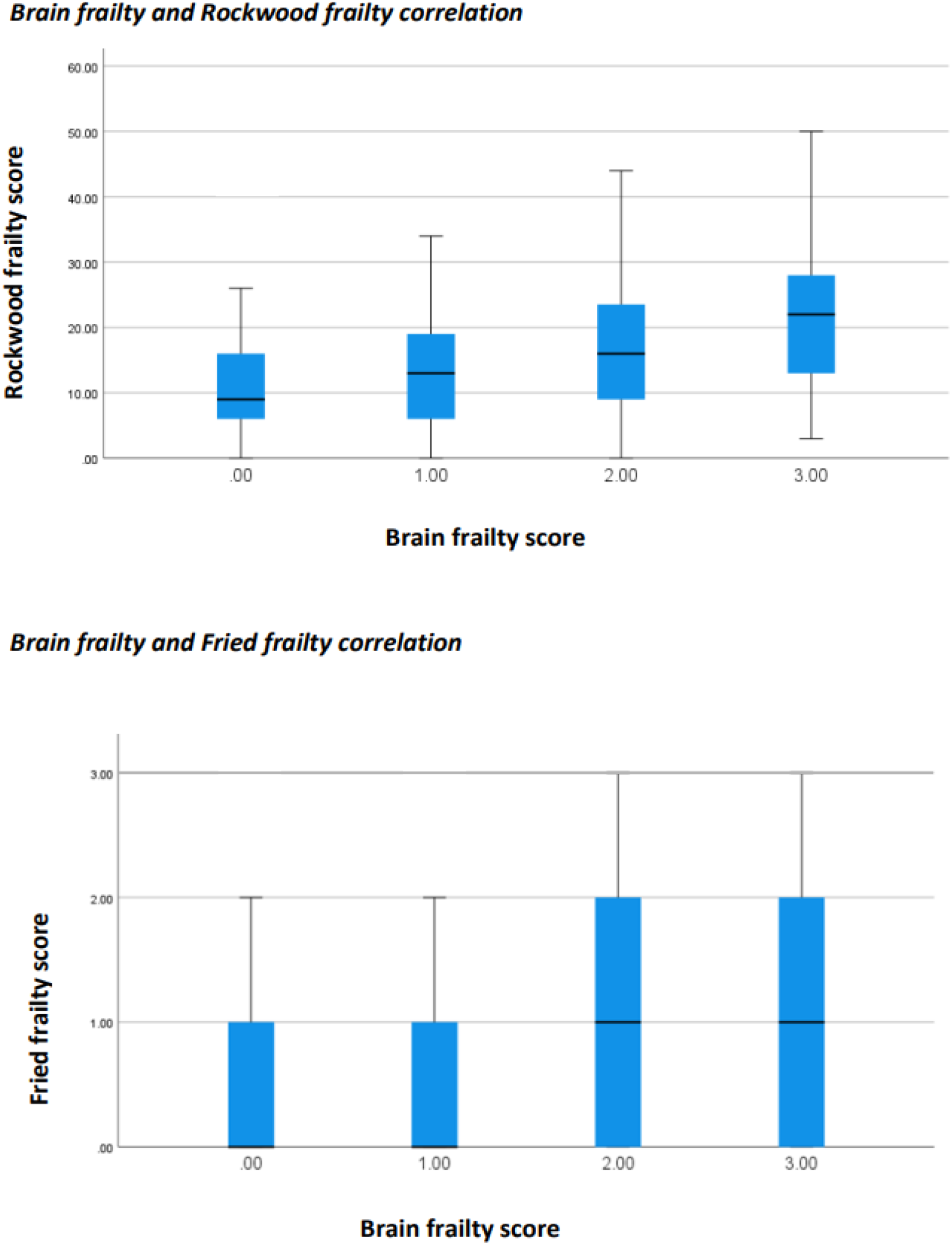
Correlation between brain frailty and traditional frailty measures

### Prevalence of brain frailty in those who are physically frail

A total of 106/332 (32%, 95%CI=27%-37%) participants had a brain frailty score of 1; 96/332 (29%, 95%CI=24%-34%) had a score of 2; and 46/332 (14%, 95%CI=10%-18%) had a score of 3.

Fifty-nine out of 330 (18%; 95%CI=14%-22%) participants were categorised as frail (cut point>0.24) on the Rockwood frailty index. Twelve out of 59 (20%; 95%CI=11%-33%) of those categorised as frail on the Rockwood frailty index had a brain frailty score of 1; 24/59 (41%; 95%CI==28%-54%) had moderate bran frailty (score=2) and 19/59 (32%, 95%CI=21%-46%) had a severe (score=3) brain frailty score. By comparison, of those considered to be pre-frail (172/330; 52%, 95%CI=47%-58%), 54/172 (31%, 95%CI=25%-39%) had a brain frailty score of 1, 52/172 (30%, 95%CI=23%-38%) had a brain frailty score of 2, and 19/172 (11%, 95%CI=7%-17%) had a brain frailty score of 3; and of those considered to be ‘robust’ (98/330; 30%, 95%CI=25%-35%) by Rockwood frailty index (score<8), 40/98 (41%, 95%CI=31%-51%) had a brain frailty score of 1, 19/98 (19%, 95%CI=12%-29%) had a brain frailty score of 2, and 7/98 (7%, 95%CI=3%-14%) had a brain frailty score of 3.

One-hundred-seventy out of 320 (53%, 95%CI=48%-58%) participants were classified as frail on the Fried frailty measure. Forty-seven out of 170 (28%, 95%CI=21%-35%) of those who were frail according to Fried frailty measure had a brain frailty score of 1; 60/170 (35%, 95%CI=28%-43%) had a brain frailty score of 2, and 30/170 (18%, 95%CI=12%-24%) had a brain frailty score of 3. By comparison, of those with no Fried frailty 57/150 (38%, 95%CI=30%-46%) had a brain frailty score of 1, 30/150 (17%, 95%CI=14%-27%) had a brain frailty score of 2, and 14/150 (9%, 95%CI=5%-15%) had a brain frailty score of 3.(**Figure 3**)

**Figure 3:**
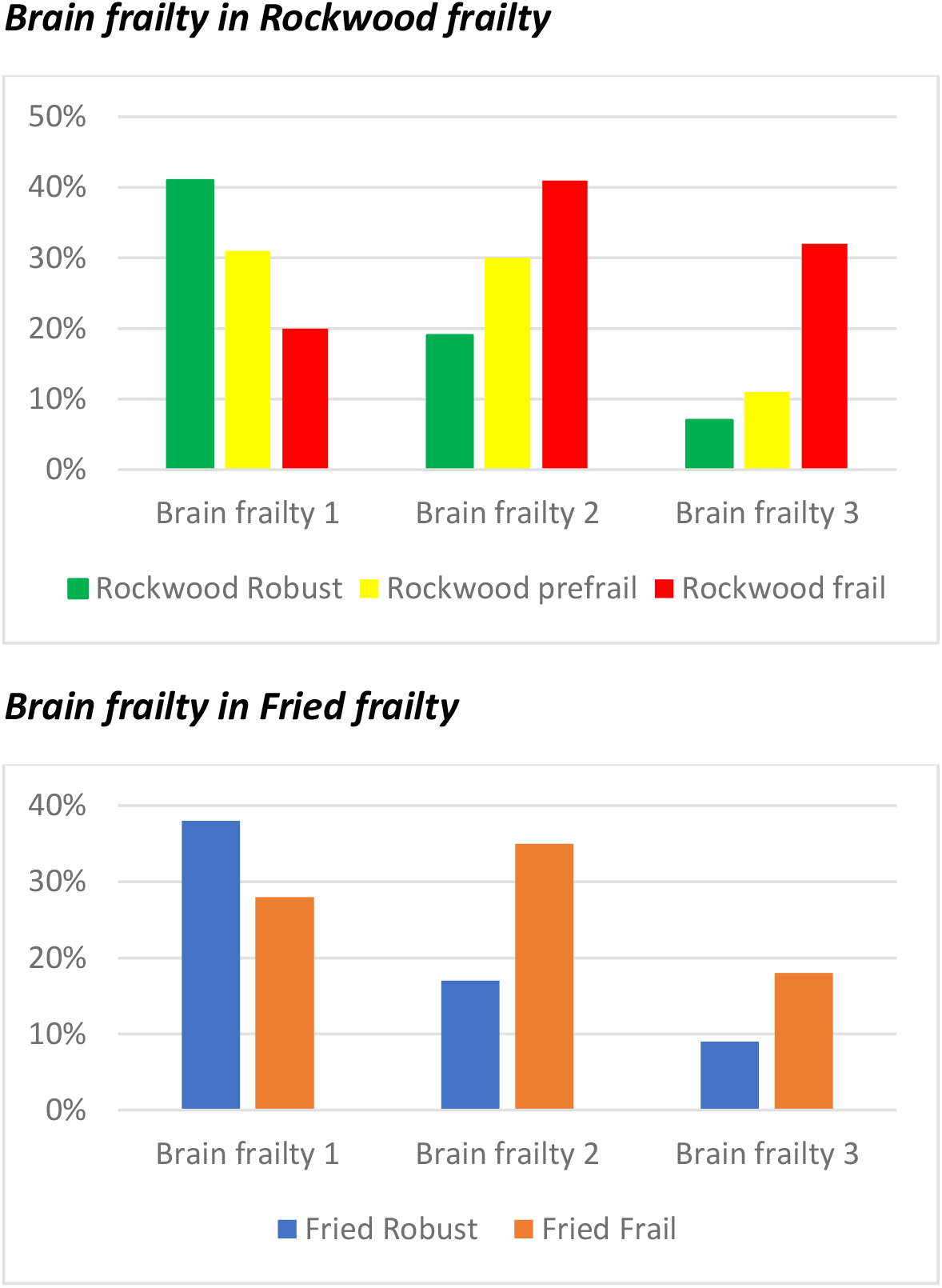
Prevalence of brain frailty according to frailty status

Overlap between measures of frailty were limited: only 22% of participants were classified as frail on both the Rockwood and Fried measures. By comparison, 41% of participants were categorised as frail on both brain frailty and Fried scale, while 17% were classified as frail on both brain frailty and Rockwood scale (17%). Degree of overlap for all three frailty measures is illustrated (**Figure 4)**.

**Figure 4:**
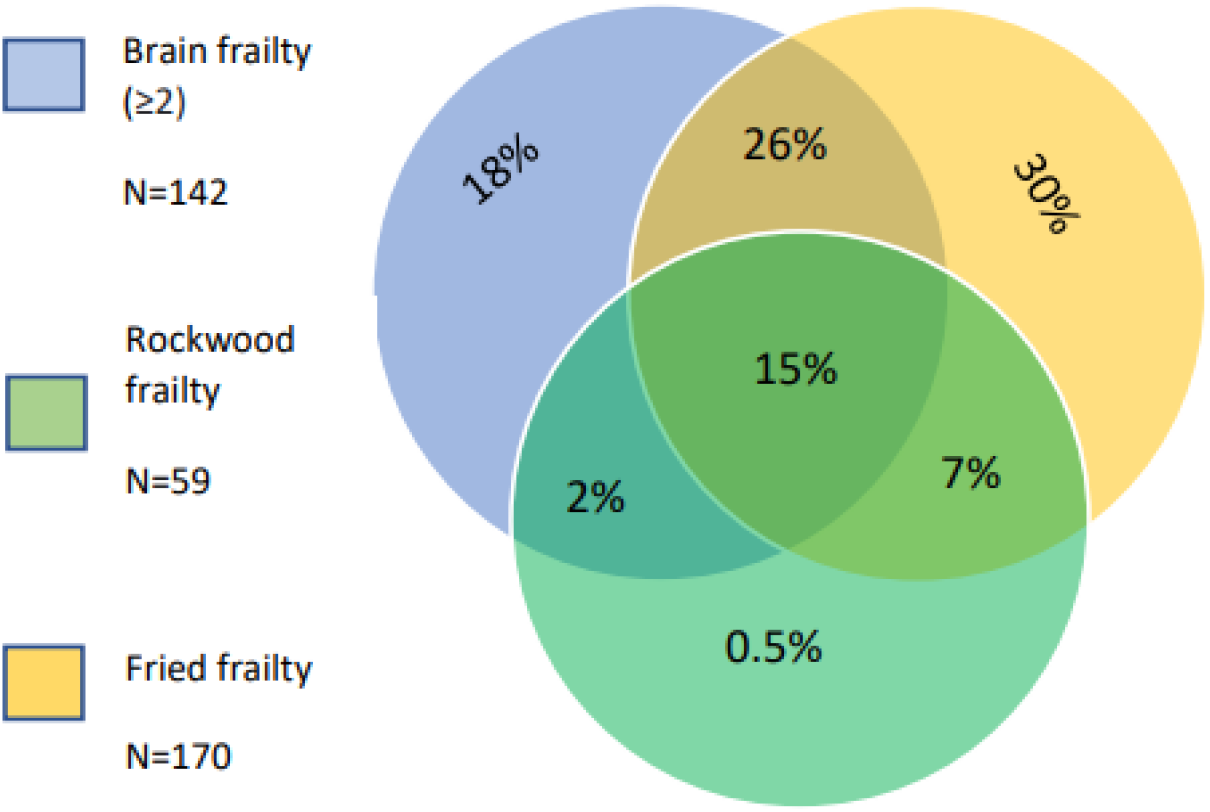
Overlap between participants categorised as frail according to each measure. ^*^Numbers are rounded to the nearest whole so % do not add up to 100%. Venn diagram depicts the percentages who were classified as having Brain frailty (scores of ≥2), Fried frailty, Rockwood frailty, or combination of each.

### Brain/physical frailty association with long-term cognitive impairment

Two-hundred-ninety-five participants were included in primary analysis; 69/295 (23.4%) had adjudicated cognitive impairment at 18 months. In Model 1, each point increase on the brain frailty scale (OR:1.64, 95%CI=1.17-2.32), Rockwood frailty index (OR:1.05, 95%CI=1.02-1.08), and Fried frailty scale (OR:1.93, 95%CI=1.39-2.67) independently increased odds of cognitive impairment 18 months after stroke. Model 2 suggested brain frailty was predictive of long-term cognitive impairment independent of both Rockwood frailty (brain frailty OR:1.47, 95%CI=1.03-2.09) and Fried frailty (brain frailty OR:1.48, 95%CI=1.04-2.12). Rockwood frailty was similarly predictive of long-term cognitive impairment independent of brain frailty (Rockwood OR:1.04, 95%CI=1.01-1.07), as was Fried frailty(Fried OR:1.78, 95%CI=1.27-2.49). In Model 3, which included adjustment for baseline cognitive status, brain frailty was no longer predictive of cognitive impairment (OR:1.40, 95%CI=0.93-2.10), while Rockwood frailty (OR: 1.04, 95%CI=1.01-1.08) and Fried frailty (OR:1.57, 95%CI=1.08-2.26) remained predictive. Model 4, which included brain and Fried or Rockwood frailty controlled for baseline cognitive score, suggested Fried frailty remained predictive of long-term cognitive impairment independent of both brain frailty and baseline AMT scores (Fried frailty OR:1.49 95%CI=1.03-2.18); however, Rockwood frailty (OR:1.04, 95%CI=0.99-1.076) and brain frailty (OR:1.29, 95%CI=0.85-1.95) were not. (All models **Table2**)

**Table 2.** Logistic regression output for models 1-4.

***Brain frailty regression output for association with 18-month cognition***
| Characteristic | Multivariable model 1 | Multivariable model 2 | Multivariable model 3 | Multivariable model 4 |
| --- | --- | --- | --- | --- |
| Age | OR:1.05; 95%CI=1.01-1.08 | OR:1.05; 95%CI=1.01-1.08 | OR:1.05; 95%CI=1.01-1.09 | OR:1.04; 95%CI=1.01-1.08 |
| Sex | OR:0.75; 95%CI=0.41-1.37 | OR:0.79; 95%CI=0.43-1.46 | OR:0.79; 95%CI=0.39-1.60 | OR:0.83; 95%CI=0.41-1.69 |
| Stroke severity | OR:1.07; 95%CI=1.00-1.15 | OR:1.05; 95%CI=0.98-1.13 | OR:1.01; 95%CI=0.91-1.14 | OR:0.99; 95%CI=0.88-1.12 |
| Education (years) | OR:0.92; 95%CI=0.82-1.03 | OR:0.93; 95%CI=0.83-1.04 | OR:0.96; 95%CI=0.86-1.08 | OR:0.97; 95%CI=0.87-1.09 |
| Brain Frailty | OR:1.64; 95%CI=1.17-2.32 | OR:1.47; 95%CI=1.03-2.09 | OR:1.40; 95%CI=0.93-2.10 | OR:1.29; 95%CI=0.85-1.95 |
| Rockwood Frailty | - | OR:1.04; 95%CI=1.01-1.07* | - | OR:1.04; 95%CI=0.99-1.08* |
| Fried Frailty | - | OR:1.78; 95%CI=1.27-2.49* | - | OR:1.49; 95%CI=1.03-2.18* |
| Baseline cognitive score | - | - | OR:0.72; 95%CI=0.63-0.81 | OR:0.71; 95%CI=0.62-0.81 |

| Characteristic | Multivariable model 1 | Multivariable model 2 | Multivariable model 3 | Multivariable model 4 |
| --- | --- | --- | --- | --- |
| Age | OR:1.06; 95%CI=1.03-1.09 | OR:1.05; 95%CI=1.01-1.08 | OR:1.06; 95%CI=1.02-1.09 | OR:1.04; 95%CI=1.01-1.08 |
| Sex | OR:0.84; 95%CI=0.46-1.53 | OR:0.79; 95%CI=0.43-1.46 | OR:0.86; 95%CI=0.43-1.72 | OR:0.83; 95%CI=0.41-1.69 |
| Stroke severity | OR:1.04; 95%CI=0.97-1.12 | OR:1.05; 95%CI=0.98-1.13 | OR:0.98; 95%CI=0.88-1.10 | OR:0.99; 95%CI=0.88-1.12 |
| Education (years) | OR:0.92; 95%CI=0.81-1.03 | OR:0.93; 95%CI=0.83-1.04 | OR:0.97; 95%CI=0.86-1.09 | OR:0.97; 95%CI=0.87-1.09 |
| Brain Frailty | - | OR:1.47; 95%CI=1.03-2.09 | - | OR:1.29; 95%CI=0.85-1.95 |
| Rockwood Frailty | OR:1.05; 95%CI=1.02-1.08 | OR:1.04; 95%CI=1.01-1.07 | OR:1.04; 95%CI=1.01-1.08 | OR:1.04; 95%CI=0.99-1.08 |
| Fried Frailty | - | - | - | - |
| Baseline cognitive score | - | - | OR:0.70; 95%CI=0.62-0.79 | OR:0.71; 95%CI=0.62-0.81 |

***Fried frailty regression output for association with 18-month cognition***
| Characteristic | Multivariable model 1 | Multivariable model 2 | Multivariable model 3 | Multivariable model 4 |
| --- | --- | --- | --- | --- |
| Age | OR:1.07; 95%CI=1.04-1.10 | OR:1.05; 95%CI=1.01-1.08 | OR:1.06; 95%CI=1.03-1.10 | OR:1.04; 95%CI=1.01-1.08 |
| Sex | OR:0.93; 95%CI=0.50-1.71 | OR:0.86; 95%CI=0.46-1.62 | OR:0.89; 95%CI=0.44-1.80 | OR:0.86; 95%CI=0.42-1.76 |
| Stroke severity | OR:1.04; 95%CI=0.96-1.12 | OR:1.04; 95%CI=0.96-1.13 | OR:0.99; 95%CI=0.88-1.11 | OR:0.99; 95%CI=0.89-1.12 |
| Education (years) | OR:0.90; 95%CI=0.80-1.02 | OR:0.92; 95%CI=0.82-1.04 | OR:0.96; 95%CI=0.85-1.08 | OR:0.97; 95%CI=0.86-1.09 |
| Brain Frailty | - | OR:1.48; 95%CI=1.04-2.12 | - | OR:1.37; 95%CI=0.91-2.07 |
| Rockwood Frailty | - | - | - | - |
| Fried Frailty | OR:1.93; 95%CI=1.39-2.67 | OR:1.78; 95%CI=1.27-2.49 | OR:1.57; 95%CI=1.08-2.26 | OR:1.49; 95%CI=1.03-2.18 |
| Baseline cognitive score | - | - | OR:0.72; 95%CI=0.63-0.82 | OR:0.73; 95%CI=0.64-0.82 |

**Model1=Age, sex, stroke severity, education controlled for as covariates; Model 2= model 1+brain frailty or Rockwood frailty or Fried frailty (dependent on comparison) controlled for as covariates; Model 3=model 1+ baseline cognitive score controlled for as covariates; Model 4=model2 plus baseline cognitive score controlled for as covariates.**

Sensitivity analysis restricted to those with 18-month cognitive test data only was fully consistent with our primary results. (**Supplementary materials S3)**

### Subgroup analyses

Thirty-five participants had a pre-existing diagnosis of dementia. Subgroup analysis excluding participants with a formal diagnosis of dementia at baseline were fully consistent with our primary results.

Forty-six participants had a TIA only. Subgroup analysis additionally excluding participants with a TIA were also consistent with our primary results.(**Supplementary materials S4)**

## Discussion

Our findings support the validity of the brain frailty scale and suggest it has a relationship with traditional measures of frailty—particularly Rockwood accumulated deficits frailty which incorporates a greater cognitive component than the physical focused Fried frailty measure. Almost ¾ of those who are frail according to the Rockwood frailty index had moderate to severe brain frailty scores and the prevalence of moderate to severe brain frailty increased as frailty status moved from robust to pre-frail to frail. However, despite the apparent association between brain frailty and Rockwood frailty, it is also clear that each of our respective frailty measures have independent value as predictors of long-term cognitive impairment following stroke. Our findings are consistent with Wallace et al^23, 24^, suggesting frailty increases brain pathology but that it also plays a role in development of dementia above and beyond the brain pathogenesis.

Although brain frailty and Rockwood frailty were no longer significant predictors of long-term cognitive impairment after adjusting for baseline cognitive test performance, up to ¾ of stroke survivors cannot fully complete traditional pen-and paper cognitive tests ^25^, hence frailty measures may be useful as alternate prognostic indicators of long-term cognitive impairment in these instances.

The Fried frailty phenotype was distinct from the two other frailty measures evaluated here in that it appears to have prognostic value independent of both imaging-based brain frailty and baseline cognitive test performance. Most stroke survivors show cognitive impairment in the immediate aftermath of stroke, but trajectories of recovery/further decline are variable. Measurement of Fried frailty may therefore be of value when evaluating cognitive prognosis. Fried frailty is also a potentially modifiable form of frailty^26^ and as such could be a target for interventions designed to minimise the long-term cognitive impact of a stroke. Physical and cognitive outcomes are often interrelated^27^; increased exercise has been associated with reduced white matter hyperintensities and cerebral atrophy in older adults,^28^ and physical activity plus risk factor reduction interventions have been shown to reduce rates of long-term cognitive decline.^29^ This suggests that long-term cognitive outcomes may be improved by addressing issues of physical function, such as frailty. However, studies aiming to treat symptoms of the Fried frailty phenotype have produced mixed results overall^30^, and while symptoms of Fried frailty may be improved through exercise or nutritional therapies, this may do little to address the root cause of the syndrome. In addition, concerns have previously been raised regarding the feasibility of screening for Fried frailty in the acute stroke setting,^1^ thus establishing when and how to measure this condition in stroke also requires further exploration.

Whether brain frailty itself is treatable remains to be established. Previous studies^31^ have suggested that antihypertensive medications may help to slow the progression of white matter hyperintensities and that this may in part play a role in reducing risk of dementia. However, evidence is limited^32^, hence this is an important avenue for further investigation.

### Strengths and limitations

Our study involved a highly inclusive stroke cohort, and we followed best practice guidance for conduct and reporting. We ensured blinding and double scoring to minimise risk of bias and conducted a series of subgroup analyses to ensure our results were translatable to clinical practice. Our population appears generalisable, and we used CT imaging data to generate brain frailty scores meaning our results will be applicable to most stroke services. Despite this, there are important limitations to note, 18-month cognitive test data were unavailable for a proportion of our sample, meaning that outcome assessment was reliant upon clinical case notes and primary/secondary care medical records. Although reliable dementia diagnoses can still be achieved using this approach,^33^ some minor cases of cognitive impairment may have been missed; however, our sensitivity analysis suggests that this limitation did not significantly impact upon our results.

There were differences between risk of missing data and risk of in-study death based on pre-stroke frailty/brain frailty status. These differences may have impacted study significance values and effect sizes. However, this limitation is synonymous with longitudinal frailty and cognition research and may not be surmountable. Our post hoc analyses were not powered to include the additional variables investigated; hence, the lack of significant predictive power of Rockwood frailty and brain frailty after including additional covariates could be related to lack of adequate statistical power.

## Conclusions

Evaluating the combination of pre-existing leukoaraiosis, atrophy and old infarcts via routinely collected CT scan images appears to be a valid method of establishing a person’s brain resilience or vulnerability following stroke. Many stroke survivors who show signs of frailty before their stroke also show signs of brain frailty. However, despite an apparent co-occurrence between brain frailty and traditional frailty, each respective measure retains independent value for predicting long-term adverse cognitive outcomes. Our findings emphasise the public health importance of life-course risk factors on the development of dementia, ^34^ and suggest physical frailty may be a viable target for interventions aiming to reduce post-stroke dementia risk.

## Data Availability

All data is avaialable upon reasonable request from the corresponding author.

## Acknowledgements

We would like to thank the Scottish Neurological Research Fund, Stroke Association, and Chief Scientist Office of Scotland for supporting this work.

